# Half Year Longitudinal Seroprevalence of SARS-CoV-2-antibodies and Rule Compliance in German Hospital Employees

**DOI:** 10.1101/2021.03.29.21254538

**Authors:** Jonas Herzberg, Tanja Vollmer, Bastian Fischer, Heiko Becher, Ann-Kristin Becker, Hany Sahly, Human Honarpisheh, Salman Yousuf Guraya, Tim Strate, Cornelius Knabbe

## Abstract

**Introduction:** COVID-19, caused by SARS-CoV-2, is an occupational health risk especially for healthcare employees. This study was designed to determine the longitudinal seroprevalence of specific immunglobolin-G (IgG)-antibodies in employees in a hospital setting.

**Methods:** All employees including healthcare and non-healthcare workers in a secondary care hospital were invited to participate in this single-center study. After an initial screening, a 6 months follow-up was done which included serological examination for SARS-CoV-2-IgG-antibodies and a questionnaire for self-reported symptoms, self-perception and thoughts about the local and national hygiene and pandemic plans.

**Results:** The seroprevalence of SARS-CoV-2-IgG-antibodies was 0.74% among 406 hospital employees (95% confidence interval) (0.75% in healthcare workers, 0.72% in non-healthcare workers), initially recruited in April 2020, in their follow-up blood specimen in October 2020.

In this study, 30.54% of the participants reported using the official German corona mobile application and the majority were content with the local and national rules in relation to Coronavirus restrictions.

**Discussion:** At the 6 months follow-up, the 0.74% seroprevalence was below the reported seroprevalence of 1.35% in the general German population. The prevalence in healthcare workers in direct patient care compared with those without direct patient contact did not differ significantly.

## Introduction

Severe acute respiratory syndrome coronavirus type 2 (SARS-CoV-2), causing the corona-virus-disease-19 (COVID-19) has spread from China throughout the whole world, beginning in Autumn 2019.[1] With a rising number of infected patients in hospitals and especially in Intensive Care Units, the medical staff became the essential cogs of health care systems around the world.[2] Protecting these employees is still one of the most important duties within this crisis to keep global health care systems intact and functional.[3] As SARS-CoV-2 is transmitted by droplet infection,[4] undiagnosed infections in medical staff members can lead to an uncontrolled person-to-person spread to other healthcare workers (HCW) and patients causing a breakdown of the health care system.[5]

In addition to viral detection using polymerase chain reaction (PCR) and now upcoming rapid antigen tests[6] used in detection of current infection, serological tests for SARS-CoV-2-specific antibodies are another option especially to retrospectively detect asymptomatic or oligosymptomatic infected persons within a defined time period.[7] Studies showed a high rate of seroconversion for immunoglobulin G (IgG) within two to three weeks after disease onset.[8–11] The longevity of the specific antibodies still is under discussion[12] and studies showed no seroconversion for initial PCR-positive tested individuals.[13]

The cost-effective and easy performance of the antibody-tests leads to an increasing amount of studies describing the seroprevalence of SARS-CoV-2-antibodies in defined groups around the world.[14–20] Especially within the group of HCW, this tool is frequently used to detect asymptomatic infected individuals, especially following the thought of a potential protection after a serological immune response.

To evaluate the longitudinal seroprevalence in a secondary care mid-sized hospital, a prospective trial was initiated. All employees such as cleaning staff, housekeeping and administration staff in addition to HCW were included. As in the first phase of the trial all inhabitants of an affiliated convent were included in the study, as they lived adjacent and were at times involved in aspects of non-direct patient care. This enabled a diverse age range of study participants, allowing a good comparison with the general population. During the study period, the national protection plan limited private meetings to 10 persons, required wearing face masks in public transportation and limited public events following social distancing of at least 1.5m, restaurants and shops remained open.

## Methods

### Study design

The “Prospective Sero-epidemiological Evaluation of SARS-CoV-2 among Health Care Workers” (ProCoV-study) is an ongoing longitudinal trial.[20] The secondary care hospital is located in the province of Schleswig-Holstein near the border of the city of Hamburg. During the entire study period of 6 months (14th of April till 20th of October), 36 PCR-confirmed COVID-19-patients were treated on isolation wards and in the intensive care unit at the study center.

Within the first study period starting in April 2020 all hospital employees and nuns between 18 and 90 years were given the opportunity to participate in this trial. As in the first phase of the study, no pretesting was performed. After the first 9-week-longitudinal evaluation of seroprevalence and PCR-positivity within this cohort, a mid-term-evaluation of the seroprevalence was performed at 6 months, that also included a questionnaire that evaluated thoughts on national hygiene regulatory and travel-/social-restrictions.

Written and informed consent was given by all study participants prior to enrolment.

### Study activities

During the first phase of the trial, all participants completed an initial questionnaire with items on demographics, general health and medication, primary working area and risk for potential SARS-CoV-2 exposure. Participants were asked to provide a weekly oropharyngeal swab and a weekly blood specimen. Results of this phase were published in October 2020.[20]

6 months after the start of this trial, all participants were re-invited to provide a follow-up blood specimen and a follow-up questionnaire including questions about the estimated personal risk for COVID-19, potential symptoms and the satisfaction with the local and national protection protocols during October 2020.

At the beginning of the study period in April 2020 a strict local hygiene protocol was established including basic hygiene standards like wearing hospital clothing and surgical masks. Personal protective equipment (PPE), including filtering face piece masks type 2 or 3 (FFP-2/FFP-3) was used routinely by employees working with suspected or confirmed COVID-19 patients. To reduce possible contacts, restrictions for visitors were enforced during the period.

The antibody-testing was performed using the semiquantitative anti-SARS-CoV-2-ELISA (IgG) from Euroimmun (Lübeck, Germany) detecting the S1 domain of the SARS-CoV-2 spike-protein with, according to the manufacturer, a specificity of 99.0% and sensitivity of 93.8% after day 20 of infection.[21] All positive and equivocally positive results were verified using two different SARS-CoV-2-ELISA (IgG): one detecting the viral nucleocapsid using the Architect SARS-CoV-2 IgG (Abbott, Wiesbaden, Germany) and the second the LIAISON SARS-CoV-2 S1/S2 IgG assay (DiaSorin Deutschland GmbH, Dietzenbach, Germany) which detects the S1- and S2 domain of the viral spike protein.

### Statistical analysis

Data were analyzed using IBM SPSS Statistics Version 25 (IBM Co., Armonk, NY, USA). All variables are presented as means or medians with standard deviation. Categorical variables are shown as numbers with percentages. Fisher’s exact test or chi-square test was used to determine relationships between categorical variables depending on size of groups. Exact 95% confidence intervals were provided where appropriate. Differences between groups were analyzed using t-tests and with logistic regression models to adjust for sex and age differences between groups. A p-value < 0.05 was considered statistically significant.

### Ethical approval

After approval by the Ethics Committee of the Medical Association Schleswig-Holstein, this trial was registered with the German Clinical Trial Register (DRKS00021270). All study activities were conducted in accordance with the Declaration of Helsinki.

## Results

In the initial study period, there were 871 participants. After calling for a 6 months follow-up, 406 of the initial participants with a median age of 44.18 years handed in a completed questionnaire and an additional blood sample for serological testing (follow-up-rate: 46.61%). This follow-up-cohort included 268 HCW and 138 non-HCW. 76.6% of the participants were female (**Table 1**). The participants in the follow-up did not differ.

**Table 1:** Comparison of characteristics of enrolled hospital employees by working area (Healthcare vs. non-healthcare). ^a^ Chi-Square Test ^b^ Fisher’s exact test ^c^ t-Test SD: standard deviation; BMI: body mass index; PCR: polymerase chain reaction

| Characteristics | HCW (n = 268) | Non-HCW (n = 138) | p-value |
| --- | --- | --- | --- |
| Median age (mean ± SD) | 41.38 ± 12.09 | 49.66 ± 15.17 | **<0.001<sup>c</sup>** |
| Female Sex, n (%) | 191 (71.27) | 120 (86.96) | **<0.001<sup>a</sup>** |
| BMI (mean ± SD) | 25.80 ± 5.71 | 26.11 ± 6.87 | 0.624<sup>c</sup> |
| Chronic medical condition, n (%) | 109 (40.67) | 72 (52.17) | **0.027<sup>a</sup>** |
| Cardiac | 39 (14.55) | 33 (23.91) | **0.019<sup>a</sup>** |
| Pulmonary | 23 (8.58) | 15 (10.87) | 0.454<sup>b</sup> |
| Metabolic | 40 (14.93) | 23 (16.67) | 0.646<sup>a</sup> |
| Immunologic | 14 (5.22) | 7 (5.07) | 1.000<sup>b</sup> |
| Other | 42 (15.67) | 26 (18.84) | 0.418<sup>a</sup> |
| Current smoker, n (%) | 70 (26.12) | 28 (20.29) | 0.194<sup>a</sup> |
| Specific symptoms, n (%) | 119 (44.40) | 40 (28.99) | **0.003<sup>a</sup>** |
| Cough | 52 (19.40) | 7 (5.07) | **<0.0001<sup>a</sup>** |
| Fever | 19 (7.09) | 4 (2.90) | 0.112<sup>b</sup> |
| Sore throat | 66 (24.63) | 23 (16.67) | 0.066<sup>a</sup> |
| Sniff | 73 (27.24) | 29 (21.01) | 0.171<sup>a</sup> |
| Headaches | 56 (20.90) | 24 (17.39) | 0.400<sup>a</sup> |
| Fatigue | 49 (18.28) | 15 (10.87) | 0.052<sup>a</sup> |
| Taste distortion | 3 (1.12) | 2 (1.45) | 1.000<sup>b</sup> |
| Anosmia | 3 (1.12) | 2 (1.45) | 1.000<sup>b</sup> |
| Use of official warning mobile application, n (%) | 92 (34.33) | 32 (23.19) | **0.021<sup>a</sup>** |
| Quarantine, n (%) | 33 (12.31) | 13 (9.42) | 0.414<sup>b</sup> |
| Reported positive PCR test, n (%) | 5 (1.87) | 3 (2.17) | 1.000<sup>b</sup> |
| Positive anti-SARS-CoV-2-IgG antibodies | 2 (0.75) | 1 (0.72) | 1.000<sup>b</sup> |

There was a significant difference in the reported presence of typical COVID19-symptoms between the HCW and non-HCW-group, where the HCW presented more often with typical symptoms, especially cough (19.40% vs. 5.07%, p-value <0.001). After adjustment for sex and age in a logistic model, the effect was not significant (p=0.071). There was no significant correlation between typical symptoms and positive PCR-test (6 out of 159 participants with symptoms, p-value 0.061) and between symptoms and seroconversion (2 out of 159 participants, p-value 0.564). Even potentially high specific symptoms such as anosmia or taste distortion (each 1 out of 5 participants reported this symptom, p-value 0.095) showed no significant correlation to positive PCR-test within the study population.

The overall rate of self-reported ever PCR positive participants was 1.97% (95% confidence interval 0.85%-3.9%) The rates did not differ between groups (1.87% in HCW vs. 2.17% in non-HCW). The rates of positive anti-SARS-CoV-2-IgG antibodies were 0.74 in total group (95% CI 0.15% - 2.2%) The percentages also did not differ between the groups (0.75% vs. 0.72% in non-HCW).

8 participants (1.97%) reported a previous positive SARS-CoV-2-PCR test, while only 3 participants showed a positivity for anti-SARS-CoV-2 IgG-antibodies at the 6 months follow-up mark. Following this evaluation, 6 participants with a previous positive SARS-CoV-2-PCR test did not show a positive antibody-test. One participant with a positive antibody-result reported no previously positive PCR-test result (**Table 2**). Not all participants reporting to have had a positive PCR reported any kind of symptoms whereas 2 of the 3 participants with a positive antibody-status reported distinct symptoms (**Table 2**).

**Table 2:** Correlation of positive PCR-test, anti-SARS-CoV-2-IgG-antibodies and symptoms. Euroimmun assay (equivocal: ratio ≥ 0.8 to < 1.1, seropositive: ratio ≥ 1.1)

| **Participant** | **Reported positive PCR** | **Initial Euroimmun IgG ratio** | **Follow-up Euroimmun IgG ratio** | **Reported symptoms** |
| --- | --- | --- | --- | --- |
| 1 | Yes | 3.4 | 2.9 | Long-term cough, headaches, fatigue |
| 2 | Yes | 0.1 | 0.1 | No |
| 3 | Yes | 0.2 | 0.4 | No |
| 4 | Yes | 0.1 | 0.2 | fever, fatigue, sore throat, limb pain |
| 5 | Yes | 0.2 | 0.2 | fatigue, sore throat, limb pain |
| 6 | No | 0.6 | 1.2 | No |
| 7 | Yes | 0.1 | 0.1 | cough, headaches, fatigue |
| 8 | Yes | 1.3 | 1.0 | fever |
| 9 | Yes | - | 3.2 | headaches, fatigue, sore throat, limb pain, taste distortion, anosmia |

In the non-HCW follow-up-cohort the majority listed prior comorbidities, with a significant difference to the HCW-group (40.67% in HCW, 52.17% in non-HCW, p-value 0.027) in the univariate analysis. Especially the area of cardiac comorbidities differed significantly between both groups (14.55% vs. 23.91% in non-HCW, p-value 0.019). The effect remained significant in the logistic regression analysis (p=0.012). The difference of body mass index between both groups was also significant (p<0.001). There was no significant difference in smoking behaviors between both groups. In a logistic model adjusted for sex, age, BMI and smoking the difference between the groups disappeared.

In the HCW the usage of the official mobile warning Application was significantly higher than in the non HCW group (34.33% vs. 23.19% in non-HCW, p-value 0.023 after adjustment for age and sex in a logistic model).

The personal risk was estimated to be low or less in 68.7% of all participants without significant differences between both groups (68.8% in Non-HCW, 68.7% in HCW).

The rates of participants requiring a period of quarantine did not differ significantly between HCW and non-HCW. The main reason for quarantine in HCW was due to professional contact to positive tested patients or co-workers (22/34) whereas the reasons for isolation in non-HCW was distributed into professional contact (7/14), private contact (2/14) and return from holidays in high-risk regions (5/14).

After 6 months of strict hygiene protocols within the hospital and with rapidly changing national pandemic regulations, 36.4% of the participants were content or very content with the national regulations (**Figure 1**) and 63.8% reported to be very content or at least content with the local in-hospital protocols (**Figure 1**).

**Figure 1:**
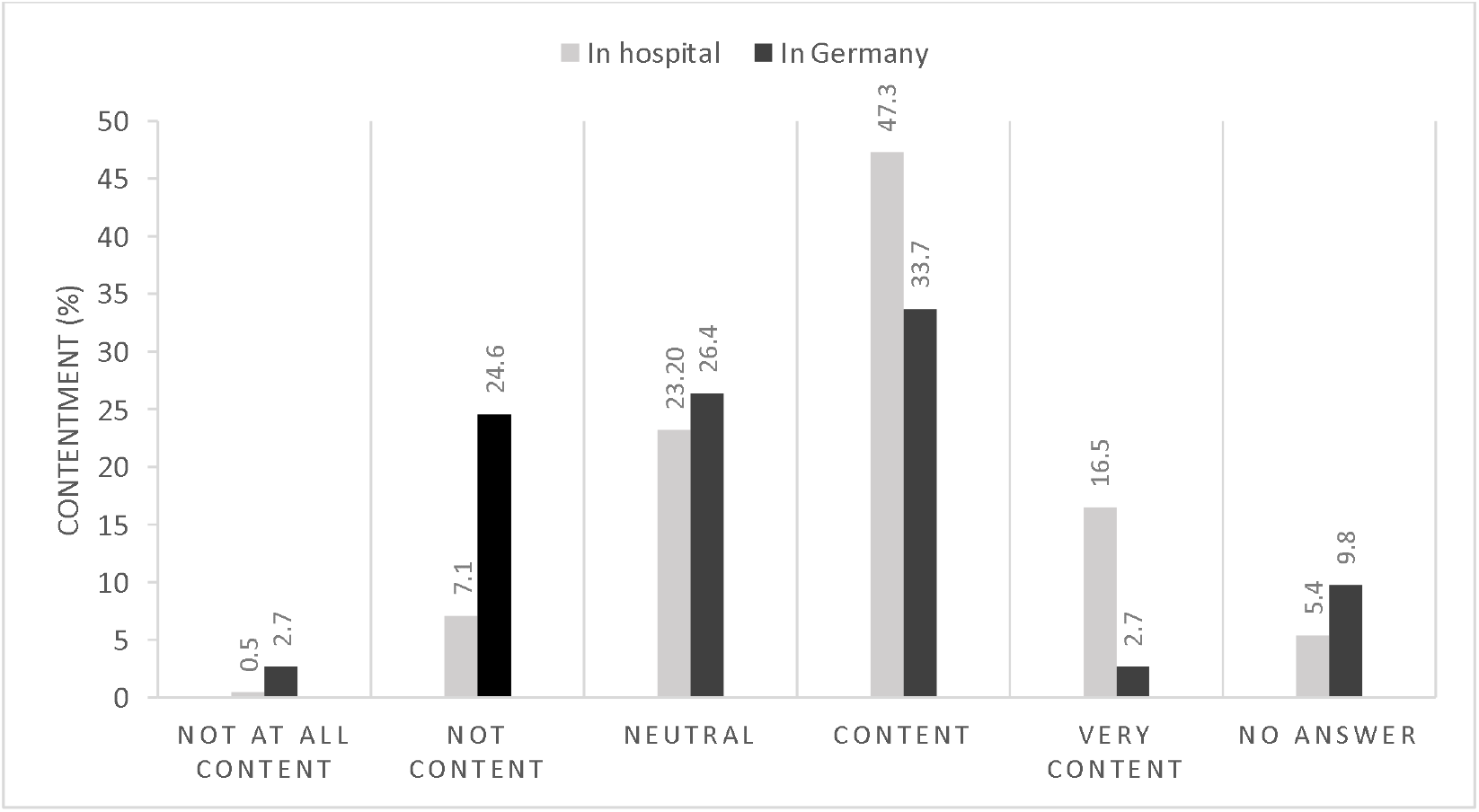
Contentment (%) with local (grey) and national (black) pandemic protocol, n=406).

Asked for the causes of their discontent with the national reglementation 87.39% described them as to loose whereas only 5.41% declare the restrictions in Germany as to strict. At local hospital level one participant described the rules as to strict (**Figure 2**)

**Figure 2:**
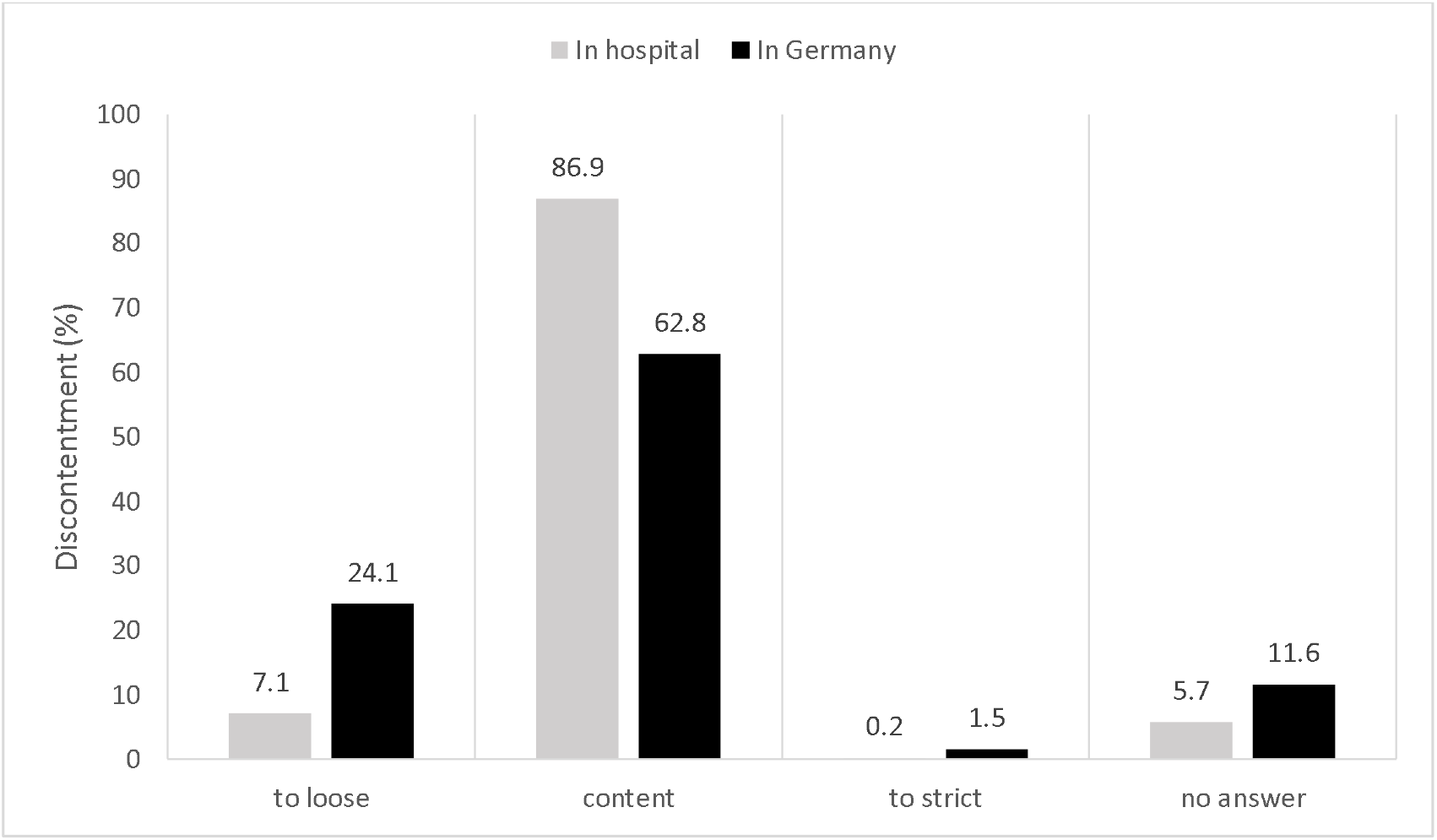
Causes for discontentment (%) with local (grey) and national (black) pandemic protocol, n=406). Neutral, content and very content are summarized under “content” in this figure.

Almost half of the participants reported that they adhered to the rules completely (HCW: 42.2%, non-HCW: 57.2%) (**Figure 3**).

**Figure 3:**
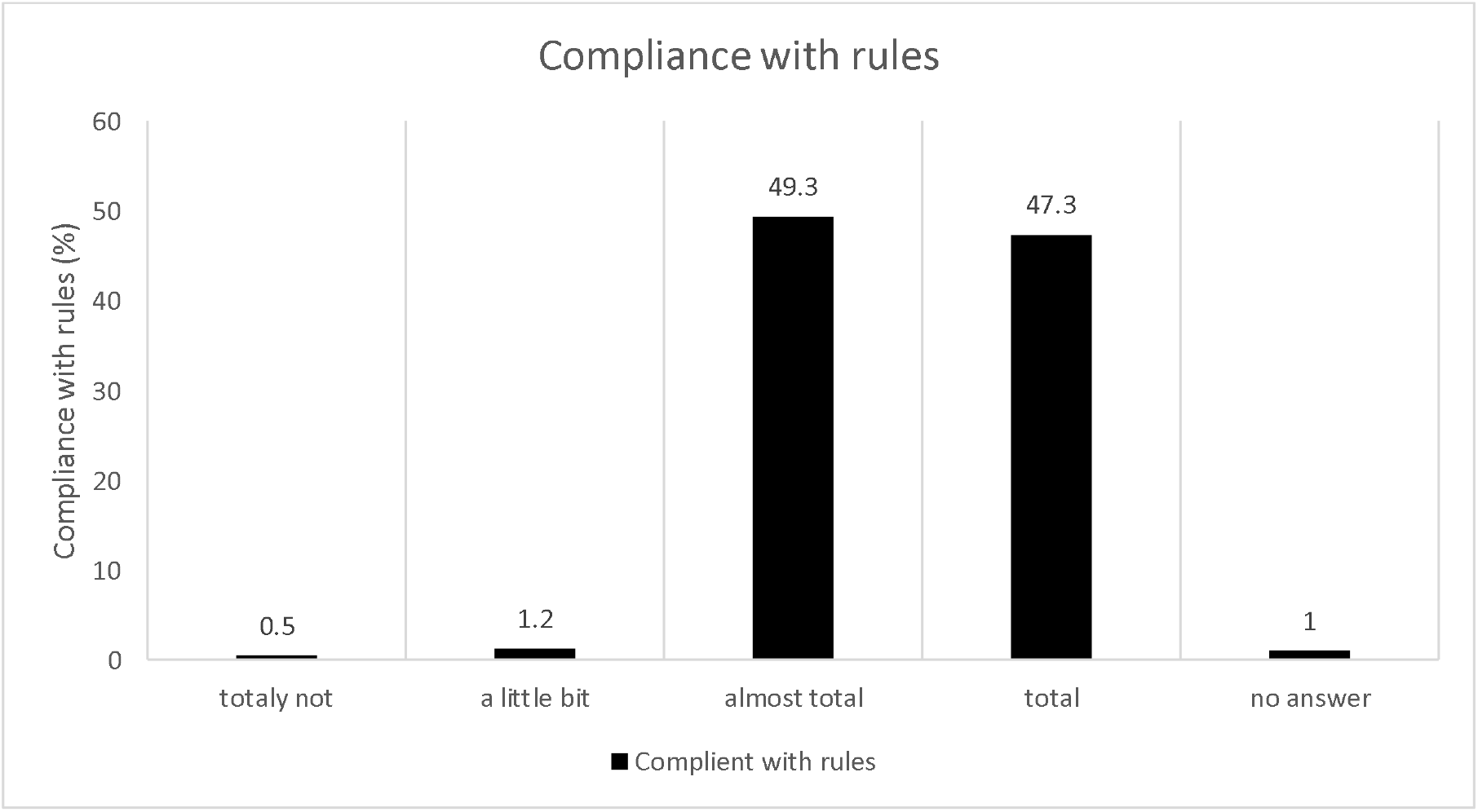
Employees self-reporting compliance with rules (%), n=406.

Adding all participants reporting to follow the rules almost completely (HCW: 54.1%, non-HCW: 39.9%) there is no difference between both groups (96.3% in HCW vs. 97.1% in non-HCW).

## Discussion

This study provides one of the first 6-month follow-up reports on the occurrence of specific anti-SARS-CoV-2-IgG antibodies within a high-risk group of hospital employees caring for COVID19 patients. Within the follow-up period, 3 participants (0.74%) showed a seroconversion. 8 participants (1.97%) reportedly had a positive throat swab prior to the blood sample being taken.

### Seroprevalence

In this trial not all participants who reportedly had a positive PCR showed specific antibodies within the follow-up blood specimen. This could be caused by an early decrease of antibody-titer as Ibarrondo et al. and Long et. al showed in their work.[12,22] Furthermore different studies suggested, that asymptomatic or oligosymotimatic infected patients do not seroconvert.[21,23,24] Tan et al. showed in their analysis that patients with a higher disease severity are more likely to develop a stronger antibody response as we saw in two participants with high IgG-ratio in our study (**Table 2)**.[25]

The overall seroprevalence of anti-SARS-CoV-2-IgG-antibodies in this trial is comparable to data from other German hospitals ranging from 0% in a 5-day-setting[14] up to > 12% in a larger longitudinal study reported by Malfertheimer et al.[16] Longitudinal data reporting the conversion to seropositive status are limited so far. Behrens et al. showed in their CoCo-trial a longitudinal prevalence of anti-SARS-CoV-2-IgG-antibodies of 1.86% within 6 weeks.[26] Comparing with our initial data from the initial 9 week phase of the study, we had shown a seroprevalence of 4.36% (with the limitation of different assays used in that evaluation).[20] The data presented here is comparable with the seroprevalence in the normal German population of around 0.91% between March and May 2020.[27] These rates are significantly lower than data reported by a population-based trial from Switzerland reporting a seroprevalence up to 10.6% in the same time period.[28] Following the large seroprevalence-study conducted by the Robert-Koch-Institute in blood donors, the seroprevalence within the German population is 1.35% in almost 50,000 tested blood samples.[29]

As the seroprevalence correlates with the local infection rate, data from other regions around the world are difficult to compare. **Table 3** provides an overview of seroprevalence studies amongst health care providers in European countries.

**Table 3:** Seroprevalence of anti-SARS-CoV-2-IgGantibodies in different European countries.

|  | **Country** | **Design** | **Participants** | **Seroprevalence of anti-SARS-CoV-2-IgG-antibodies** |
| --- | --- | --- | --- | --- |
| Krankenhaus Reinbek | Germany | single-center | 406 | 0.74% |
| Steensels et al.[30] | Belgium | single-center | 3056 | 10.6% |
| Garcia-Basteiro et al.[31] | Spain | single-center | 578 | 9.3% |
| Jespersen et al.[32] | Denmark | multi-center | 17971 | 3.4% |
| Sotgiu et al.[33] | Italy | single-center | 202 | 14.4% |
| Rudberg et al.[34] | Sweden | single-center | 2149 | 19.1% |

Due to limitations in the study we were unable to assess possible infection routes, in order to evaluate if the prior infection was work-acquired or due to community exposures to SARS-CoV-2. Paderno et al. showed infection routes in HCW.[35]

### Self-perception and evaluation

Almost 40% of all participants reported symptoms suspicious for a potential SARS-CoV-2-infection, such as fever or cough. Due to the low PCR-positivity rate and seroconversion, there is no significant correlation between these symptoms and a possible SARS-CoV-2-infection.

In contrast to the research presented by Behrens et al., the majority of the participants in this study estimated their own personal risk for a prior infection as low or very low independent of their working area.[7]

In the cohort presented here, 30.54% of the participants used the official mobile application “Corona-Warn-App” presented by the Robert-Koch-Institute.[36] There was a significantly higher user rate in the HCW-group. This might be due to a younger mean age in this group. As this application uses a decentralized system without data collection, the only known data is the number of downloads instead of actual users. According to the federal German government the application was downloaded 20 million times by the 20^th^ of October 2020.[37] Comparing this with the amount of smartphone uses in Germany, which was 58 million in 2019[38], the user rate in the society is around 34%, comparable with the rates of our study individuals.

Looking to the rate of quarantine for different reasons 10.8% is a relevant proportion for a healthcare provider (12.31% in HCW vs. 9.42 in non-HCW). This in combination with a high rate of employees presenting specific symptoms lead to the conclusion that a protocol for screening using PCR or rapid antigen-tests is needed preventing the breakdown of a healthcare system within a global pandemic.[39]

In addition to the serological follow-up-examination, the questionnaire included some questions about the perception of the local and global pandemic protocols. The data showed that the majority of study participants abided by the rules almost completely or else completely without differences between both groups (non-HCW: 97.1%, HCW:96.2%). Only a few participants declared being unable to abide by the rules. This is an important factor, as even a few individuals could jeopardize the whole hospital and in addition the whole health care system.

The majority of the participants were content with the local hygiene protocols and pandemic plan and no participtant described it as to loose, whereas a quarter were not content with the national hygiene protocols and pandemic plans. In this group just 5.41% rate the national rules as to strict.

### Limitation

The major limitation of this study is its single-center structure. Furthermore, the participant rate of 46.66% of all initially included participants for the follow-up, is relatively low. This might be caused by the effect of shift work, holidays or participants no longer working in the study center and lost to follow-up. Females are highly overrepresented in both groups, representing a common trend in health care workers.[40] Due to the low number of seroconverted participants within this 6 months follow-up, the study has a low power detecting characteristics of seroconverted participants encourage such a seroconversion. The only possibility for grouping the participants, is according to their profession and not their working area, as a large number of employees work in multiple areas of the hospital (both high and low risk areas). This may be compounded by the fact that in the study center there were no completely separated pathways for possible COVID-19 patients

## Conclusion

The data presented provides one of the first longitudinal sero-epidemiological assessments of the prevalence of specific antibodies against SARS-CoV-2 in healthcare workers compared with hospital employees without direct patient care.

Moreover, this study presents the self-perception of hospital employees with regards to their views of the local and national protection protocols, by individuals intimately involved in the fight against this global pandemic.

## Data Availability

The datasets generated and analysed during the current study are not publicly available but are available from the corresponding author on reasonable request.

## Acknowledgments

We’d like to acknowledge the effort of all team members at the Krankenhaus Reinbek St. Adolf-Stift and at all participating laboratories that analyzed the samples in addition to their daily workload.

Special thanks to Rebecca Zimmer (linguistic enrichment) for her intense help and input throughout the entire process of this study.

